# Bilingualism measures in children: A critical review of content overlap, development, and pragmatic quality

**DOI:** 10.1101/2024.10.30.24316440

**Authors:** Kai Ian Leung, Monika Molnar

**Affiliations:** Department of Speech-Language Pathology, University of Toronto, Toronto, CANADA; Rehabilitation Sciences Institute, University of Toronto, Toronto, CANADA

**Keywords:** bilingual, child, questionnaire, overlap, pragmatic quality, measure development

## Abstract

**Purpose:** A variety of assessment tools (e.g., questionnaires) measure the type and degree of bilingualism in children in both research and clinical settings. Although these tools are often assumed to evaluate the same constructs and be interchangeable, this may not be the case, as indicated by other recent reviews. This review critically evaluated existing measures of child bilingualism, focusing on item-content overlap, measure development, and pragmatic quality.

**Method:** A database and manual search identified studies on child bilingualism measure development, which were then appraised using the Psychometric and Pragmatic Evidence Rating Scale and the Consensus-based Standards for the Selection of Health Measurement Instruments (COSMIN).

**Result:** Analysis across the six identified measures showed weak between-measure content overlap, with less than one quarter of items shared on average, suggesting they assess different constructs.

Ratings indicated varied pragmatic quality, especially in assessor burden (training, interpretation). COSMIN evaluations also highlighted shortcomings in measure design and development.

**Conclusion:** The findings underscore the need for improved content validity and better pragmatic criteria for the clinical use of these tools. We offer recommendations for measure selection dependent on use case (e.g., setting-specific needs) and suggestions for future bilingualism measure development, prioritizing a pragmatic approach.

---

Despite the emphasis on integrating research evidence into clinical practice, there is a clear shortage of robust studies concerning bilingual client assessment and intervention. Most of the existing research consists of observational and case studies conducted across varied bilingual populations (Thordardottir, 2010). A major challenge in evaluating this body of bilingual research lies in the inconsistency in how bilingualism is defined, measured, and reported across research (e.g., Williams et al., 2021). This lack of standardization complicates drawing clear conclusions, both for clinical applications and for theory development for that matter. To properly interpret research and effectively inform clinical practice, it is crucial that language experiences are consistently measured and reported using reliable tools. Equally important is the need for clinicians to carefully consider a client’s full language history when providing care, as research has shown the importance of language-related barriers in accessing services, quality of care, and health outcomes (Bowen, 2001). For researchers and clinicians alike, the selection of valid, reliable tools that capture a comprehensive language history is vital - not only to ensure the validity and interpretability of research findings, but also to support effective clinical decision-making.

It is typically assumed in clinical research and practice that all language background questionnaires are designed to evaluate the same concepts and would provide the same outcome and information about a participant or client. Because of this, one could choose any of the available questionnaires – or even, one could design their own set of questions to that effect. However, it has been demonstrated that questionnaires intended for bilingual adults, do not tend to evaluate the same constructs and not necessarily interchangeable. In their recent study, Dass et al., (2024) employed the content overlap analysis method to classify and compare the item content of adult questionnaires measuring bilingualism. They categorized questionnaire items into five global categories relevant to bilingualism: production, switching, exposure, subjective statements, identity, and history/acquisition. Their analysis revealed minimal overlap among the included questionnaires, suggesting that each questionnaire they examined address distinct aspects of the bilingual experience – hence they would likely not produce the same description of a bilingual individual.

A variety of assessment tools (e.g., questionnaires) are available to estimate the type and degree of bilingualism in children as well. While they are used in both research and clinical practice, these are primarily designed for a research audience (Anderson et al., 2018; Li et al., 2006; Marian et al., 2007; Paradis, 2011; Paradis et al., 2010). In a comprehensive review and comparability analysis of 48 such questionnaires, Kašćelan and colleagues (2022) identified as many as 32 constructs—broad conceptual categories such as language exposure, proficiency, and language use, which were included in these questionnaires—and 194 components, or the specific subcategories or items used to measure these constructs. Just like the review of the adult questionnaires, their analysis revealed a lack of standardization in how these constructs and components are operationalized, making it difficult to cross-compare results or apply them interchangeably in research and practice. For instance, the construct of language *exposure* (i.e., quantity and quality of interactions a child may have with a given language) was operationalized using 15 different components, each measuring a different aspect of exposure, such as time spent with speakers of each language, the number of conversational partners, or the variety of language environments. Users of these questionnaires must carefully consider specific constructs and components of each questionnaire to ensure appropriate use and interpretation of its findings.

In addition to an instrument’s content, its psychometric and pragmatic characteristics are important aspects. In the case of language background questionnaires, these have been not yet examined. All assessments used in clinical and research settings, including language background questionnaires, should be pragmatic – important to stakeholders, actionable, brief, and sensitive to change – to maximize utility (Glasgow & Riley, 2013; Kroenke et al., 2015). Psychometric properties of assessment tools are also an important factor. The Consensus-Based Standards for the Selection of Health Measurement Instruments (COSMIN; Mokkink et al., 2010) identifies three key domains: validity, which ensures the tool measures what it is supposed to (including content validity, the most important aspect, as well as construct and criterion validity); reliability, which refers to the tool’s consistency over time (including internal consistency, test-retest reliability, and measurement error); and responsiveness, which ensures the tool can detect changes in the measured construct over time, validating any change in scores. While traditional tool development has prioritized psychometric considerations, a pragmatic measure places equal or greater emphasis on other practical, pragmatic criteria, such as feasibility and low user burden in real word settings (Glasgow, 2013). A pragmatic approach is particularly well-aligned with clinical fields like speech-language pathology, where tools and interventions must be not only valid and reliable but also effective in routine clinical practice (Schliep et al., 2017). Despite this, no reviews to date have explicitly adopted a pragmatic approach to assessment evaluation, highlighting the need for more attention for its potential in speech-language practice.

## The Current Study

Although numerous language questionnaires exist, few have been developed with both psychometric and pragmatic considerations in mind. In a review, Kašćelan et al. (2022) emphasized the need for a critical approach to documenting bilingualism, particularly in terms of questionnaire validity, administration, and suitability for practice. To address this gap, our study specifically sought to critically review child language background measures. Unlike previous reviews (e.g., Kašćelan et al., 2022), we focused on published measures only because they are publicly available to clinicians and they offer more of the necessary data to assess pragmatic quality and psychometric properties, particularly content validity – the extent to which a measure represents the construct it aims to assess.

In addition, we adapted the methodology used in Dass et al. (2024) to provide a systematic method for evaluating the overlap of existing bilingualism measures for children, with specific details provided for each measure. This approach enables an overlap comparison across all measures, highlights critical gaps and ultimately can aid in the selection of appropriate tools for bilingual assessment. In contrast to Kašćelan et al. (2022), whose comparability analysis was thematic to a subset of their identified constructs and components, our study employs conservative categorizations derived from the literature and presents results for each measure individually, allowing for clearer, more specific comparisons. As well, our approach provides a structured analysis and greater clinical applicability.

Finally, our study includes a pragmatic appraisal to ensure the assessment tools are not only valid but also have potential for implementation in clinical settings. This pragmatic focus, which has not been fully explored in prior reviews, evaluates the extent to which current tools are actionable and effective in clinical practice as well as for research.

In the current study, our goals were to: (i) identify relevant measures of child bilingualism; (ii) evaluate the overlap (commonalities and differences) between these measures; and (iii) appraise the measure development and pragmatic quality to better understand its clinical utility.

## Methods

### Measure identification

We used a multi-pronged search strategy to identify studies describing the development or validity of a bilingualism measure for paediatric populations. This approach included (i) studies identified from a previous review; (ii) an updated database search; and (iii) an informal manual search. Kašćelan and colleagues’ (2022) review previously identified 81 measures were in their review of questionnaires quantifying the bilingual experience in children. To update their search, we used the same keywords (Supplementary Materials), applying their search strategy across two databases, APA PsycINFO, and OVID Embase + Embase Classic. Additionally, we performed a manual informal search using Google Scholar. The searches were last updated in February 2024.

Eligibility criteria were adapted from Kašćelan et al. (2022) and Dass et al. (2024). Included measures were in the English language, mentioned bilingualism, were used for the evaluation of children (aged 0-18) and were the main subject of the peer-reviewed publication (e.g., describing development, testing reliability/validity). Our inclusion of English language measures was to ensure we appraised measures that were accessible both to the broader clinical audience and to the language abilities of our team. Measures were excluded if they (1) were designed for quantification of bilingualism in adults only; (2) were not focused on language; (3) were language-specific only (e.g., designed for use with a specific group of bilinguals only or with non-English bilinguals); (4) primarily concerned foreign language learning in educational settings; (5) primarily concerned speech and language disorders; (6) were duplicates or earlier iterations of a measure; and (7) could not be accessed (e.g., not openly available).

### Content overlap analysis

Following Fried’s (2017) approach, we conducted a content overlap analysis approach comprised of a multi-step process. This included: i) categorizing question items from the identified child bilingualism measures, ii) consolidating and reducing items within each measure, and iii) performing between-measure comparisons using Jaccard similarity indices to assess content similarity.

Question-item categorization followed a structured and iterative process, involving a detailed examination of each question item and review of the relevant literature. First, all question items were compiled, with sub-questions treated as separate items. The items were then sorted into categories based on key language background concepts (e.g., age of acquisition, language exposure) frequently discussed in developmental research. Categories were deductively derived from existing literature on bilingualism, with particular attention given to constructs critical for assessing bilingual abilities in both languages. Following Wall & Lee (2022), we adopted a conservative strategy to avoid overestimating heterogeneity in content overlap. Ambiguous items were flagged for further review and resolved through discussion and consensus among authors. Items could be attributed to multiple categories, although this was rare. The categorization process underwent several rounds of re-evaluation, incorporating an inductive approach for misfitting items, until all inconsistencies were resolved. Although comparisons with other categorization systems could provide additional insight, we chose to remain within the established framework to avoid overexpanding categories and inflating variation. This approach allowed us to conservatively estimate overlap and maintain a clear focus on the key bilingualism constructs relevant to each measure.

After the initial categorization, we did a within-measure review of the material to identify questions that may be redundant, following the method outlined by Fried (2017). Questions that were worded similarly or in reverse were grouped as a single item. Duplicate questions intended for different caregivers or languages were also consolidated into one item.

Next, an analysis of between-measure content was performed. Using the established list of (sub)categories, we organized all questions from each measure into a master spreadsheet to determine the frequency of each category’s appearance in each measure (Supplementary Materials). Unlike Dass et al. (2024), we opted not to separate demographics and language-related items in the analysis, to demonstrate a comprehensive and practical analysis of measure content. A category table was used to produce a matrix, with items coded as follows: “1” if featured in the scale and “0” if not featured.

Jaccard similarity indices were calculated for each measure pair, by determining the ratio of shared question items to the sum of unique and shared items across the two measures. Jaccard index strength was interpreted following Fried (2017), ranging from very weak (0.00–0.19), weak (0.20–0.39), moderate (0.40–0.59), strong (0.60–0.79), and very strong (0.80–1.0). Average overlap for each measure and overall average overlap across all measures was calculated. To explore the effect of measure characteristics on overlap, a correlation analysis using each average overlap with the measure length (the number of categories covered by the scale) and with the percentage of idiosyncratic (unique) items present in the measure. We adapted the script from Fried (2020, see article for the corrigendum specific OSF link) for the statistical analysis in R software (R version 4.3.3, 2024).

### Quality appraisal

Pragmatic criteria scores from Psychometric and Pragmatic Evidence Rating Scale (PAPERS) were used across identified measures (Lewis et al., 2021; Stanick et al., 2021). The criteria evaluated by PAPERS include factors such as the cost of the measure, its length, language readability, and the burden on assessors in terms of training and interpretation. Each criterion is rated on a 6-point scale (i.e., −1 - poor to 4 - excellent). Higher scores are desired, as they indicate a more pragmatic measure.

The Consensus-based Standards for the Selection of Health Measurement Instruments (COSMIN; Mokkink et al., 2018; Terwee et al., 2018) checklist was employed to assess the content validity and methodological quality of the development of the included measures. Part 1a focuses on standards for evaluating the quality of research conducted to identify relevant items for a new measure. The quality of the concept elicitation study provides information on the relevance and comprehensiveness of the items in a measure. Part 1b pertains to standards for assessing the quality of a cognitive interview study or pilot tests (e.g., surveys) performed to evaluate the comprehensiveness and comprehensibility of the measure. Each standard is rated on a 4-point scale (e.g., very good, adequate, doubtful, inadequate; 0-not-applicable), with the overall rating obtained by taking the lowest rating of any of the standards (“worst score counts” method).

## Results

### Goal 1: Identify measures

As shown in Figure 1, the database and informal searches yielded 370 papers, in addition to the 81 measures initially identified in Kašćelan et al. (2022)’s review. Six measures met inclusion criteria, all targeting the caregiver/parent as informant for the measures (see Table 1 for measure characteristics). These measures included the Alberta Language and Development Questionnaire (ALDeQ; Paradis et al., 2010), Bilingual Language Questionnaire (BIPAQ; Abutbul-Oz & Armon-Lotem, 2022), Language Exposure Assessment Tool (LEAT; DeAnda et al., 2016), Language Mixing Scale Questionnaire (LMSQ; Byers-Heinlein, 2013), Multilingual Approach to Parent Language Estimates (MAPLE; Byers-Heinlein et al., 2020) and the Practitioner recommended modules of the Quantifying Bilingual EXperience^1^ (Q-BEx; De Cat et al., 2023).

**Figure 1.**
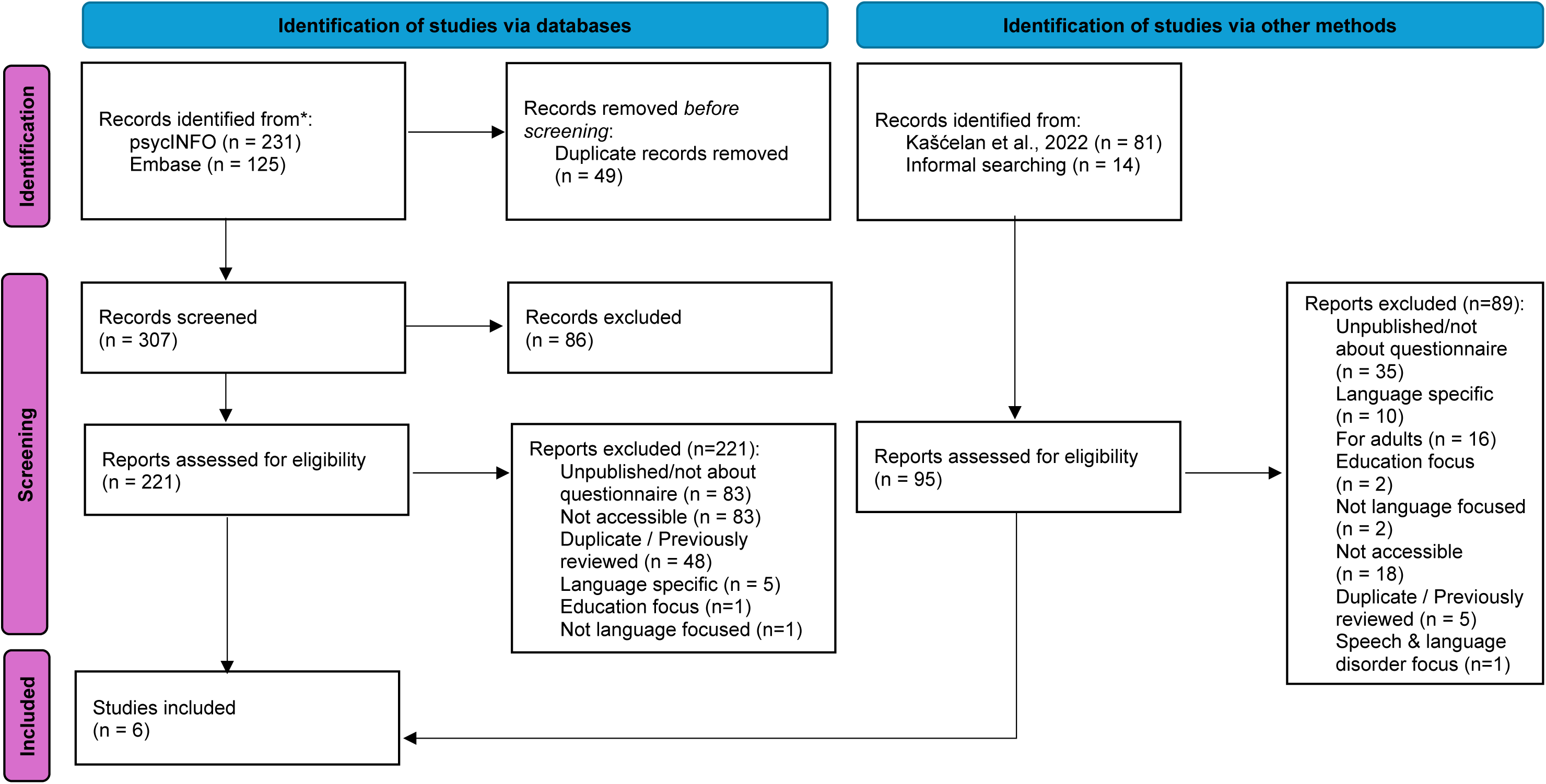
Preferred Reporting Items for Systematic reviews and Meta-Analyses (PRISMA) Flowchart showing literature identification, screening, and inclusion of the literature search.

**Table 1.**
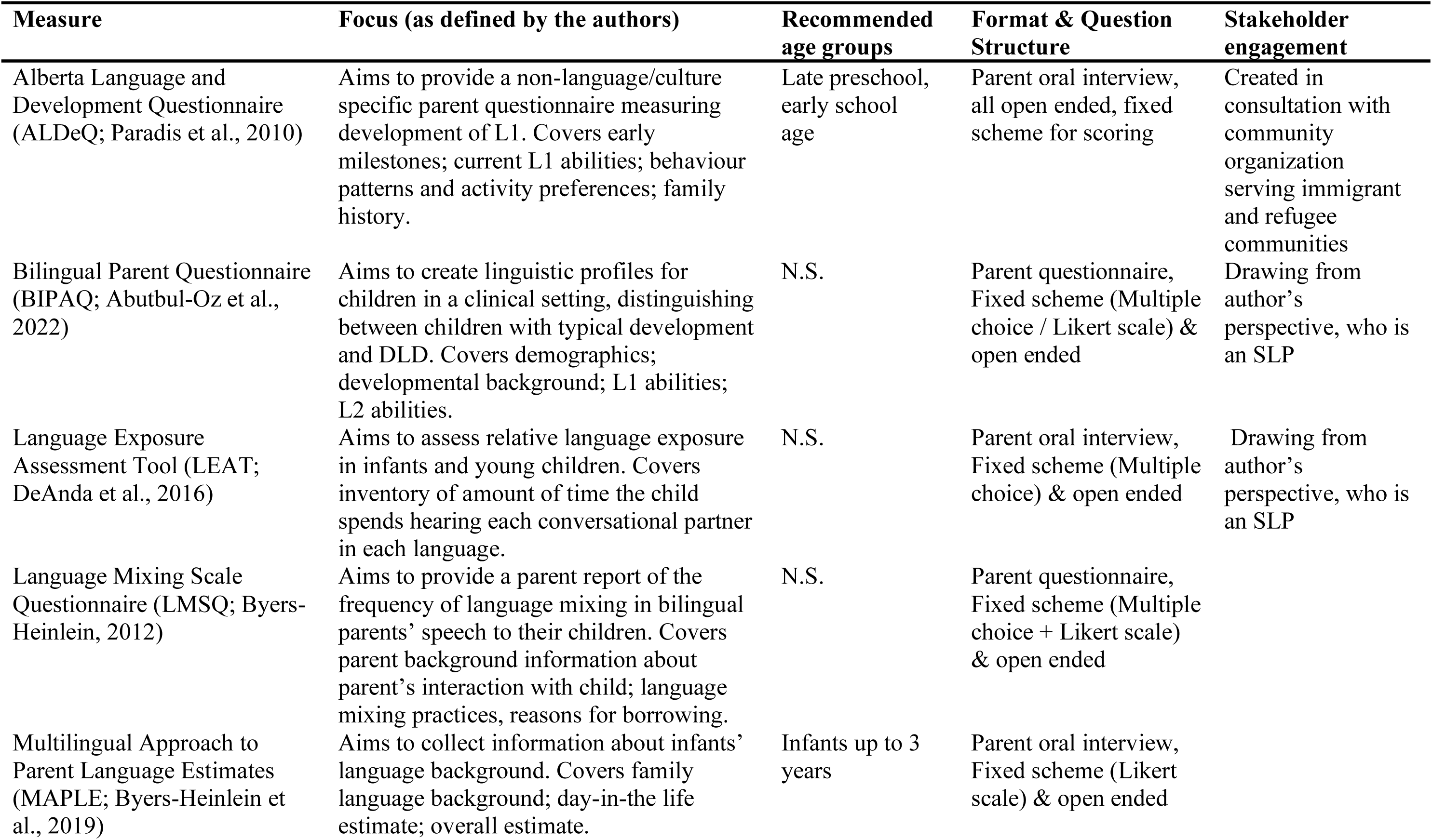
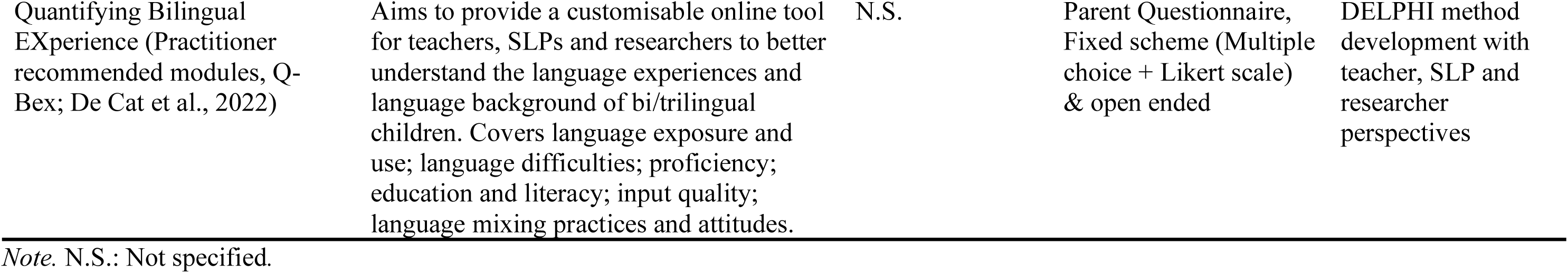
Characteristics of included measures including focus, age group, format and question types and stakeholder engagement.

| **Measure** | **Focus (as defined by the authors)** | **Recommended age groups** | **Format & Question Structure** | **Stakeholder engagement** |
| --- | --- | --- | --- | --- |
| Alberta Language and Development Questionnaire (ALDeQ; Paradis et al., 2010) | Aims to provide a non-language/culture specific parent questionnaire measuring development of L1. Covers early milestones; current L1 abilities; behaviour patterns and activity preferences; family history. | Late preschool, early school age | Parent oral interview, all open ended, fixed scheme for scoring | Created in consultation with community organization serving immigrant and refugee communities |
| Bilingual Parent Questionnaire (BIPAQ; Abutbul-Oz et al., 2022) | Aims to create linguistic profiles for children in a clinical setting, distinguishing between children with typical development and DLD. Covers demographics; developmental background; L1 abilities; L2 abilities. | N.S. | Parent questionnaire, Fixed scheme (Multiple choice / Likert scale) & open ended | Drawing from author's perspective, who is an SLP |
| Language Exposure Assessment Tool (LEAT; DeAnda et al., 2016) | Aims to assess relative language exposure in infants and young children. Covers inventory of amount of time the child spends hearing each conversational partner in each language. | N.S. | Parent oral interview, Fixed scheme (Multiple choice) & open ended | Drawing from author's perspective, who is an SLP |
| Language Mixing Scale Questionnaire (LMSQ; Byers-Heinlein, 2012) | Aims to provide a parent report of the frequency of language mixing in bilingual parents' speech to their children. Covers parent background information about parent's interaction with child; language mixing practices, reasons for borrowing. | N.S. | Parent questionnaire, Fixed scheme (Multiple choice + Likert scale) & open ended |  |
| Multilingual Approach to Parent Language Estimates (MAPLE; Byers-Heinlein et al., 2019) | Aims to collect information about infants' language background. Covers family language background; day-in-the life estimate; overall estimate. | Infants up to 3 years | Parent oral interview, Fixed scheme (Likert scale) & open ended |  |
| Quantifying Bilingual<br>EXperience (Practitioner<br>recommended modules, Q-Bex; De Cat et al., 2022) | Aims to provide a customisable online tool<br>for teachers, SLPs and researchers to better<br>understand the language experiences and<br>language background of bi/trilingual<br>children. Covers language exposure and<br>use; language difficulties; proficiency;<br>education and literacy; input quality;<br>language mixing practices and attitudes. | N.S. | Parent Questionnaire,<br>Fixed scheme (Multiple<br>choice + Likert scale)<br>& open ended | DELPHI method<br>development with<br>teacher, SLP and<br>researcher<br>perspectives |

*Note.* N.S.: Not specified.

None of the measures above were included in Dass et al.’s (2024) content analysis as they focused exclusively on adult measures. The ALDeQ, BIPAQ, LEAT, LMSQ and MAPLE were previously identified and included in Kašćelan et al’s (2022) comprehensive review; however, the Q-BEx is a newly developed tool by the same team behind the review (De Cat et al., 2023).

### Goal 2: Commonalities and differences between content of measures

#### Measure (sub)categories

Content overlap analysis was conducted across 181 question items across the six measures. The measure categorization process produced eight categories (*Age of acquisition, Interlocutor, Context/Amount, Proficiency, Demographics & Identity, Health History and Other*), which were further divided into 19 subcategories; see Table 2 for a description of these (sub)categories. The item reduction process reduced the LEAT from 23 items to 17, the LMSQ from 14 to 11, the MAPLE from 30 to 25, and the BIPAQ from 51 to 35, resulting in an overall reduction of total question items from 181 to 151. Figure 3 and Table 3 show how frequently each measure captured a specific (sub)category.

**Figure 2.**
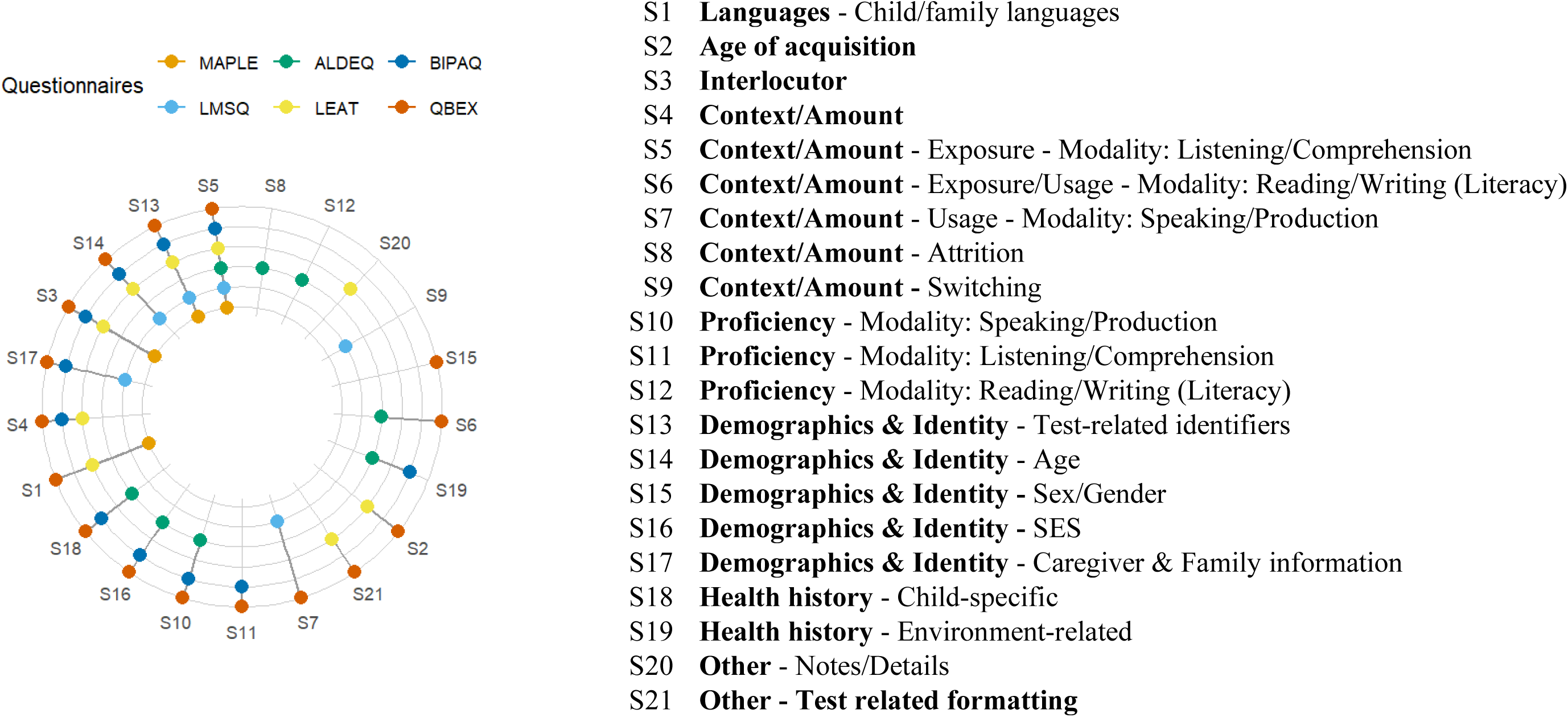
Radar chart of the (sub)category coverage for identified child measures.

**Figure 3.**
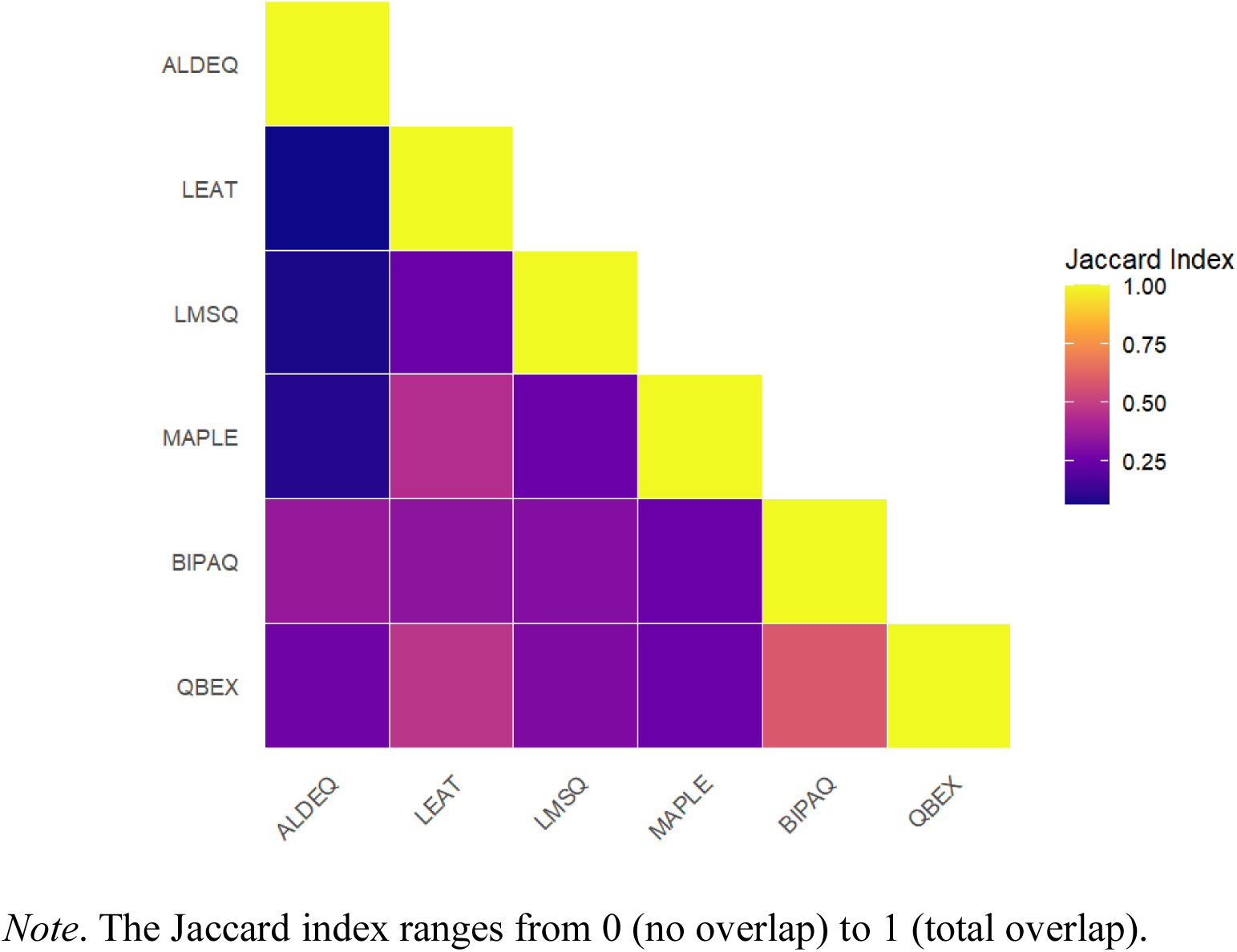
Heatmap of Jaccard similarity indices for each identified measure pair.

**Table 2.** Definitions of the categories and subcategories in the content overlap analysis.

| Category (n=8) | Subcategory (n=19) | Definitions |
| --- | --- | --- |
| Languages | Child / family languages | List of languages/dialects the child/family speak or understand |
| Age of acquisition |  | When the child first began to learn or acquire a particular language |
| Interlocutor |  | Facets of the interlocutor's language environment (e.g., nationality, immigration status, accent, native-ness, etc) |
| Context/Amount |  | Considers contextual factors/activities unrelated to a specific language modality (e.g., the child's country of birth). |
| Context/Amount | Exposure - Modality:<br>Listening/Comprehension | Considers contextual factors/activities related to language exposure through listening/comprehension |
| Context/Amount | Exposure/Usage - Modality:<br>Reading/Writing (Literacy) | Considers contextual factors/activities related to language exposure and usage through reading/writing (literacy) |
| Context/Amount | Usage - Modality:<br>Speaking/Production | Considers contextual factors/activities related to language exposure through speaking/production |
| Context/Amount | Attrition | Considers contextual factors/activities related to language attrition |
| Context/Amount | Switching | Considers contextual factors/activities related to code-switching, borrowing, mixing |
| Proficiency | Modality: Speaking/Production | Child's ability to communicate express themselves verbally effectively |
| Proficiency | Modality:<br>Listening/Comprehension | Child's ability to understand spoken language or other forms of auditory input effectively |
| Proficiency | Modality: Reading/Writing<br>(Literacy) | Child's ability to understand written text and to produce written language effectively |
| Demographics & Identity | Test-related identifiers | Information to attribute responses to child/family in the context of testing (e.g., child's name, date of test, child's identifier numbers etc.) |
| Demographics & Identity | Age | Child's age at time of test (e.g., months, years, date of birth) |
| Demographics & Identity | Sex/Gender | Child's sex/gender identity |
| Demographics & Identity | Socioeconomic status (SES) | Evaluation of family's socio-economic position (e.g., parent's education, parent's occupation, household income) |
| Demographics & Identity | Caregiver & Family information | Information regarding the family members (e.g., birth order, siblings, etc.) |
| Health history | Child-specific | Child related health history/risk factors (e.g., early language and motor development milestones, etc.) |
| Health history | Environment-related | Family related health history/risk factors (e.g., history of language disorder, difficulties etc.) |
| Other | Notes/Details | Area to expand on questions or share notes |
| Other | Test-related formatting | Structural components to the measure (e.g., formatting unrelated to purpose of measure) |

**Table 3.** Comparison of identified measures by (sub)category coverage, adjusted length, and idiosyncratic vs. discrete (sub)category representation.

| Measure | (Sub)categories captured | Adjusted measures length | Idiosyncratic (sub)categories (%) | Discrete (sub)categories (%) |
| --- | --- | --- | --- | --- |
| ALDeQ | 8 | 20 | 10 | 38 |
| LEAT | 9 | 17 | 0 | 43 |
| LMSQ | 6 | 11 | 5 | 29 |
| MAPLE | 4 | 25 | 0 | 19 |
| BIPAQ | 11 | 35 | 0 | 52 |
| Q-BEx | 16 | 43 | 5 | 76 |

*Note.* (Sub)categories captured, number of specific 21 (sub)categories the measure captures; *Adjusted measures length*, number of items per scale after combining similar items; *Discrete (sub)categories*, rate of (sub)categories captured out of 21.

Only one (sub)category - *Context/Amount - Exposure - Modality:*

*Listening/Comprehension* - was observed across all six measures (e.g., “In what situations do you tend to speak in L2 with your child?”). The next most reproduced (sub)categories included *Demographics & Identity - Test-related identifiers* (e.g., name of child, study ID), which appeared in 5/6 measures; *Demographics & Identity – Age* (e.g., date of birth), which appeared in 4/6 measures; and *Interlocutor* (e.g., questions directed to caregiver, When people hear you speak [language] can they guess that you speak another language?), which appeared in 4/6 measures.

Four idiosyncratic categories were identified. The least overlap was seen across *Proficiency Modality - Reading/Writing (Literacy)* (e.g., “Does your child like to read books or have books read to them?”), *Context/Amount – Switching* (e.g., “I often start a sentence in L1 and then switch to speaking L2”), *Context/Amount – Attrition* (e.g., “Do you think he/she may be losing the mother tongue in favour of English?”), and *Other - Notes/Details* (e.g., space for informant to elaborate).

Despite not being the longest measure in terms of number of items or (sub)categories captured, the ALDeQ contributed the highest percentage of idiosyncratic or unique (sub)categories. This was followed by the LMSQ and Q-BEx, the shortest (11 items) and longest measure (43 items) respectively. In addition to being the longest measure, the Q-BEx also captured the most (sub)categories (16, 73%). The remaining measures contained no idiosyncratic (sub)categories.

#### Measure overlap

The Jaccard similarity indices were used to estimate the overlap between measures. Figure 4 illustrates a heatmap for Jaccard indices across each measure pair and Table 4 shows the Jaccard indices for measure pairs and mean measure overlap. The average overlap was 0.28 (range: 0.17-0.37), which indicates weak measure item overlap.

**Figure 4.**
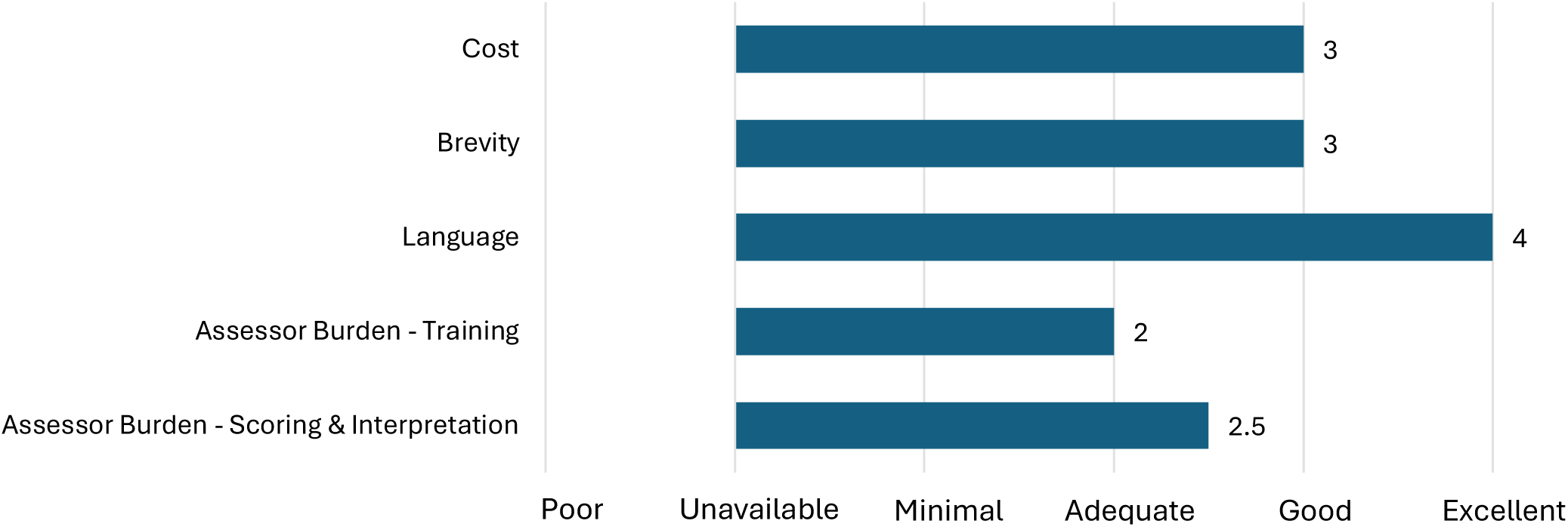
Median pragmatic quality ratings across all identified measures based on the criteria of the Psychometric and Pragmatic Evidence Rating Scale (PAPERS; Lewis et al., 2021)

**Table 4.** Pairwise overlap matrix based on Jaccard similarity indices of the identified measures.

|  | ALDeQ | LEAT | LMSQ | MAPLE | BIPAQ | Q-BEx |
| --- | --- | --- | --- | --- | --- | --- |
| ALDeQ | 1.00 | 0.06 | 0.08 | 0.09 | 0.36 | 0.26 |
| LEAT | 0.06 | 1.00 | 0.25 | 0.44 | 0.33 | 0.47 |
| LMSQ | 0.08 | 0.25 | 1.00 | 0.25 | 0.31 | 0.29 |
| MAPLE | 0.09 | 0.44 | 0.25 | 1.00 | 0.25 | 0.25 |
| BIPAQ | 0.36 | 0.33 | 0.31 | 0.25 | 1.00 | 0.59 |
| Q-BEx | 0.26 | 0.47 | 0.29 | 0.25 | 0.59 | 1.00 |
| Mean overlap | 0.17 | 0.31 | 0.24 | 0.26 | 0.37 | 0.37 |

*Note.* The Jaccard index ranges from 0 (no overlap) to 1 (total overlap).

The highest Jaccard index for any two measures was shared between the Q-BEx and BIPAQ, which had an overlap index of 0.59, indicative of moderate overlap. The ALDeQ showed the lowest individual overlap with other measures, especially with the LEAT (0.06), LMSQ (0.08) and MAPLE (0.09), which all indicate minimal content overlap.

The correlation between number of items captured and mean overlap was 0.73 (strong) suggesting that degree of measure length and overlap were related. Correlation between the percentage of measure-specific idiosyncratic items, with the mean overlap was −0.6, which suggested a moderate relationship between the proportion of symptoms uniquely probed by a particular measure and how well its content overlaps with other scales.

### Goal 3: Quality appraisal

#### Pragmatic quality of included measures

The total pragmatic rating scores for the six measures ranged from 14 to 18 (refer to Tables 5 and Figure 2 for the median rating across measures), with a median total score of 16 out of a possible 20. Most measures were readily accessible as an appendix of the journal publication and thus at low cost through journal access, with two out of six available through open access journals. Alternatively, they could be found on websites independent of the journal of publication (median score=3). Measures were concise, with a median of 26.5 question items (median score = 3), and featured highly accessible language and readability, with a median Flesch-Kincaid grade level ranging from 4th to 7th grades (median score = 4). Training information was generally not well-documented, with only two measures accompanied by manuals/training information (median score = 2). The remaining measures required some form of training/supervision for administration, but this was not well described. However, guidance on scoring and interpreting item scores was often lacking, with a median score of 2.5, especially concerning suggestions for interpreting score ranges, establishing clear cutoff scores, and addressing missing data.

**Table 5.** Pragmatic rating of included measures based on the criteria of the Psychometric and Pragmatic Evidence Rating Scale (PAPERS; Lewis et al., 2021)

|  | ALDeQ | LEAT | LMSQ | MAPLE | BIPAQ | Q-BEx |
| --- | --- | --- | --- | --- | --- | --- |
| Cost | 4 | 2 | 2 | 2 | 4 | 4 |
| Brevity (length) | 3 | 3 | 3 | 3 | 2 | 3 |
| Language | 4 | 4 | 4 | 4 | 4 | 4 |
| Assessor Burden |  |  |  |  |  |  |
| Ease of training | 2 | 3 | 2 | 2 | 2 | 3 |
| Ease of interpretation | 3 | 2 | 2 | 1 | 3 | 3 |
| Total Score | 16 | 14 | 13 | 12 | 15 | 17 |

*Note.* Scales ranges from -1 to 4 with total possible score = 20.

#### COSMIN Quality of the Measure Development

Quality of the included measures were evaluated according to Consensus-based Standards for the Selection of Health Measurement Instruments (COSMIN) standards checklist (Table 6). Given the scoring method, all measures were rated as inadequate for overall development quality. The measure design total was also mostly rated as inadequate, apart from the ALDeQ which was given the rating of doubtful. This was also the only measure to conduct a pilot study as part of its development, although the results of this was not shared. In terms of measure design, three criteria in particular were found to be lacking across all measures, particularly reporting 1) a clear origin of the construct measured (i.e., was a theory, conceptual framework or disease model used or clear rationale provided to define the construct to be measured?), 2) clear target population the measure was developed for (e.g., disease, demographic characteristics) and 3) if the measure was developed in a sample representing the target population.

**Table 6.**
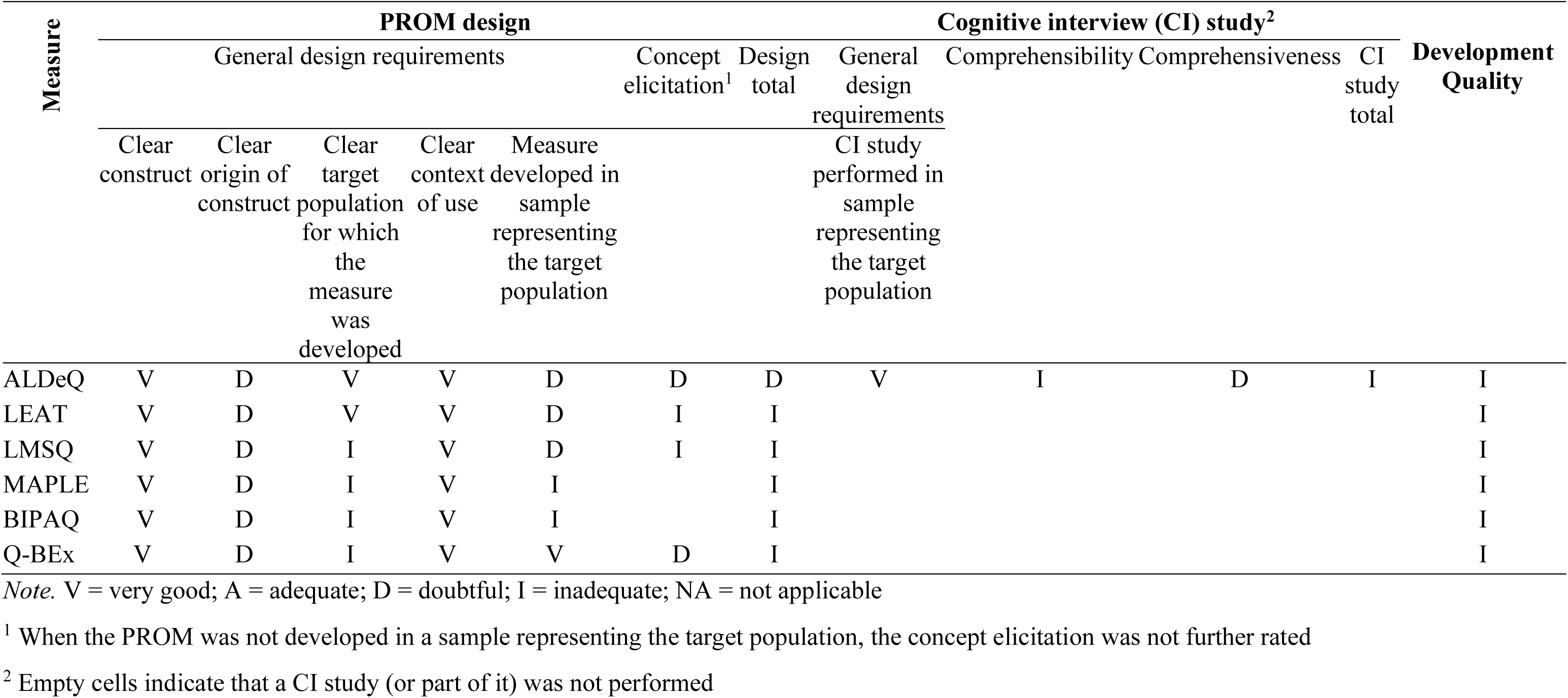
Quality of the included measure development according to COnsensus-based Standards for the selection of health Measurement INstruments (COSMIN) standards.

## Discussion

In this review, we adopted a critical approach to i) identify relevant measures of child bilingualism, ii) to provide a content overlap analysis and ii) critically review the pragmatic quality and content validity of the included measures. Our search identified only six published tools measuring child bilingualism. Our analyses of these measures covered question item content, which demonstrated limited overlap in item content. We discuss some of the more frequently observed and less frequent subcategories as well the influence of purpose on content overlap. We also observed relationships between measure length and mean content overlap, as well between the proportion of unique items and mean content overlap. Overall, the identified measures exhibited shortcomings in both development and pragmatic quality. We discuss the implications of our findings below, along with recommendations for the development of new measures and use case of existing ones.

### Content overlap among child measures

The two most frequently included subcategories relevant to categorization of language-related item content were *Context/Amount - Exposure - Modality - Listening/Comprehension* and *Interlocutor*. The *Context/Amount - Exposure - Modality - Listening/Comprehension* subcategory, present in all six measures, refers to contextual factors and activities related to language exposure through listening and comprehension. The *Interlocutor* category, included in four measures, addresses aspects of the interlocutor’s language environment, such as nationality, immigration status, accent, and native-ness. The focus on these categories highlights the importance of understanding a child’s language environment, as both the context in which exposure occurs and the characteristics of the communicative partners involved directly inform the child’s language input. Additionally, as children are still in the process of acquiring their languages, it is understandable that these measures seek to define the bilingual environment by way of language input, as opposed to output (De Houwer, 2011; Hoff and Luz Rumiche, 2011). However, it is also relevant to record in which contexts and what language is output occurring and speaking/production modality is much less featured among measures of child bilingualism (2/7 measures). In contrast, adult questionnaires placed most emphasis on language expression (*Production* – *math* and *subjective statements - speaking*), each appearing in four out of seven questionnaires (Dass et al., 2024).

Not surprisingly, other frequently addressed content in child measures included demographic-related items from *Demographics & Identity* category. These included subcategories of *Age* – the child’s age at time of test – represented in 4/6 measures and *Test-related identifiers* – information used to attribute the responses to child/family in the context of testing, such as test date, study-specific ID – in 5/6 measures. This emphasis may reflect the need to keep track of developmental progress and ensure accurate data attribution, particularly as many studies recruit families at a single time point for a cross-sectional investigation. However, recording such information also allows for potential longitudinal comparisons, enabling researchers and clinicians to track progress over time if families are followed up in later stages. This again contrasts with the demographic content overlap analysis of adult questionnaires, where age was moderately represented (3/7 questionnaires), and test-related identifiers were not considered in the analysis altogether (Dass et al., 2024). This discrepancy between child and adult measures may stem from the more stable nature of language skills in adults, hence reducing the need for age-related data. It is also important to note that test-related identifiers were omitted in both the categorization and analysis of Dass et al (2024), likely because it was not considered as a question item. Our decision to include test-related identifiers was made to reflect the practicalities of using these measures in real-world settings, where such details on identity aid in the interpretation of results and justify its inclusion.

In our analysis, categories of literacy proficiency, attrition, and language switching—all significant aspects of the bilingual experience— were not commonly addressed in the included measures. This sparse representation may be linked to the intended target audience of the measures. Notably, more than half of the measures (4/7) did not specify a recommended age group or specific audience. This is an important oversight, as knowing the appropriate age range is crucial for tailoring assessment questions. For instance, to effectively evaluate bilingual experience of school aged children, it is essential to gather information about the languages used in their schools, the second languages taught, and their literacy exposure. Therefore, here we consider and discuss the use of these measures broadly across developmental phases in children.

Literacy, encompassing the abilities to read and write, has wider, long-term implications on education and health outcomes (Office of Disease Prevention and Health Promotion, 2021; S. J. Ritchie & Bates, 2013; Stine-Morrow et al., 2015). Literacy has been part of two subcategories in our analysis, one relevant to *Context/Amount - Exposure/Use* (featured in Q-BEx and ALDeQ) and *Proficiency* (only in ALDeQ). It is possible its limited focus in the included measures may be attributable to the assumption that literacy skills are solidified after childhood as Dass and colleagues (2024) noted that literacy — albeit categorized separately across reading, writing, and other forms—was among the most frequently covered topics, with subjective statements regarding reading in particular appearing in four out of seven adult questionnaires (Dass et al., 2024). However, assessing literacy in bilingual children is crucial for early identification of gaps and can enable timely and tailored literacy interventions to support educational development.

Attrition describes the loss of proficiency in a given language over time, often observed within the context of migration for individuals experiencing first language attrition (Schmid, 2013), but may also be observed in changes to the social environments or education systems for foreign language attrition (Schmid, 2023). The existing literature on language attrition has predominantly focused on adults, which may explain its limited focus in our analysis of child measures. However, attrition has significant implications across the lifespan, and global migration contexts too are affecting children. Notably, recent figures report 28 million international child migrants, comprising 1.4% of the global child population (United Nations Department of Economic and Social Affairs [UN DESA], 2021). Adopting a developmental perspective is required to understand how attrition and language experiences evolve at different life stages (Schmitt & Sorokina, 2024).

The subcategory of switching, which we consider here together as a broader phenomenon but may sometimes be referred to separately as code-switching (intersentential) or code-mixing (intrasentential), is highly dependent on social, situational among other demands in bilinguals (W. C. Ritchie & Bhatia, 2012). Switching is a hallmark of bilingual language use and may also interact with language proficiency and dominance (Basnight-Brown & Altarriba, 2007). Across the adult questionnaires evaluated by Dass et al. (2024), language switching was similarly underrepresented, with only two questionnaires covering this aspect of bilingualism, one being the adult purpose-built Bilingualism Switching Questionnaire. The Language Mixing Scale Questionnaire (LMSQ) too was also identified as a purpose-built measure among child measures and solely featured the switching subcategory. Unlike in Dass et al., (2024), overlap did not consistently relate to the intended purpose of the measure, as seen with the LMSQ, which despite being purpose-built for language mixing and switching, did not have the least overlap with other measures. Dass and colleagues (2024) also highlighted another measure designed for a specific purpose (Bilingualism and Emotion Questionnaire) which instead had significant overlap and high item content, contrary to the other purpose-built measure. Notably, our analysis identified the LMSQ as the most concise measure among those evaluated at 11 items and effectively captured a range of categories including *Interlocutor, Context/Amount - Exposure* & *Usage* and *Demographics & Identity* despite its brevity.

Length of the measures impacted the extent of overlap among them, as demonstrated by a positive correlation. The longest measures, Q-BEx (43 items) and BIPAQ (35 items), tended to cover more subcategories and show greater overlap with each other (0.59), but not necessarily with other measures. Instead, a higher number of unique items (idiosyncratic categories) tended to correlate with less overlap, with ALDeQ containing the most unique items and being the most distinct from other measures. The ALDeQ contributed solely to items relating to (sub)categories of *Proficiency Modality - Reading/Writing (Literacy)* and *Context/Amount – Attrition* and generally tended to include broader categories across modalities (e.g., listening/comprehension, speaking production, literacy) and health history. The originality of this measure is complimented by a comprehensiveness in included dimensions, which may be related to its interest in the development of language as suggested by its name.

### Pragmatic and development appraisal

The critical appraisal using the PAPERS and COSMIN ratings revealed varied performance among the measures. Most demonstrated good pragmatic quality—being accessible, brief, and having clear, readable language—yet training documentation and guidance on scoring were often insufficient. According to COSMIN standards, development quality of the measures was found to be inadequate. It is important to note that only a few of these measures have been validated or tested for reliability and responsiveness, hence our focus here on methodological development quality. According to the COSMIN methodology, measures should demonstrate high quality in measure development (e.g., *how* the measure is designed, approach taken to item construction) and in content validity criteria, namely measure relevance, comprehensiveness and comprehensibility. However, the identified measures were found to often lack a standardized framework or consensus in their development (e.g., a clear origin for the guiding construct), and relied on the individual experiences of researchers or authors. While some of these measures were developed by clinician-researchers, the quality appraisal revealed they lacked input from other stakeholders in the form of cognitive or pilot testing with representative samples of the target population. Articles often failed to define measure characteristics, such as suitable age groups. Given that most of these measures lack psychometric validity and reliability, more steps towards improving the overall quality of measure development and its pragmatic utility should be taken.

Of note, the measures included were not all validated or tested for reliability. This constrained our ability to provide greater critical evaluation using the full COSMIN checklist. Without established psychometric properties, we cannot confidently assess how well these measures capture their intended constructs or their consistency across different populations and contexts. As a result, our findings are based on a subset of measures that may not meet the stringent COSMIN criteria, highlighting a critical gap in the current landscape of available tools.

### Considerations for measure selection

We compared the author-defined measure focus (Table 1) with our review of content overlap, development, and pragmatic quality to find alignment and divergences in certain respects. While both recognize the distinct purposes served by each measure, it is essential to consider whether the reported purposes align with the outcomes of our analyses as this can be informative in selecting the appropriate measure based on specific research or clinical goals.

The LMSQ may be preferable for research focusing on language mixing, as indicated by its purpose to provide frequency information on language mixing, item content on switching and other features (e.g., test-related identifiers, age). The LEAT and MAPLE focus on language exposure estimation for younger children and infants, which our analysis corroborated through their question items corresponding to measuring exposure in listening/comprehension, however, literacy-related modalities were absent in these measures. The Q-BEx and BIPAQ were described to have more of a clinical orientation, but we noted only the Q-BEx was able to flag concerns for early language development. The ALDeQ alone offered norms for language impairment, which align our pragmatic quality findings on its applicability in clinical settings. The Q-BEx is described as highly customizable, making it ideal for capturing nuanced bilingual language experiences in both research and clinical settings. Based on our evaluation, both the ALDeQ and Q-BEx show high pragmatic quality and greater coverage of idiosyncratic (unique) items, with the ALDeQ demonstrating higher design quality, though not without its limitations.

Certain measures are more suited for efficiency, while others are better for detail; the LMSQ and LEAT offer quick insights suitable for time-constrained contexts, whereas more comprehensive instruments like the BIPAQ and Q-BEx allow for in-depth analysis. Shorter measures may be ideal for targeted questions (e.g., LMSQ for language mixing, LEAT for exposure), while longer ones like the ALDeQ and Q-BEx provide fuller language profiles when needed and can be flexible in their context of use.

### Considerations for future measure development

Moving forward, it is recommended that questionnaires be developed with a clear framework, a transparent account of the construct’s origin and a focus on key aspects of the experience being measured. The development process should actively include input from stakeholders, such as users, and consider pragmatic criteria from the outset to support the tool’s implementation. So far, only the ALDeQ has invited community stakeholder input and the Q-BEx through its Delphi-informed development. For the Q-BEx, validation and reliability testing were still ongoing at the time of this review. The Q-BEx and ALDeQ also exhibited the highest pragmatic scores, though they also represented the highest and lowest levels of overlap, respectively.

### Study limitations

A limitation in this work may stem from the requirement for published measures in our analysis. By focusing on articles that have undergone the peer review process, we may have excluded potentially valuable instruments that are still in development or have been utilized informally in clinical practice. While this approach grounds our analysis in established research and recognized methodologies, it may not capture the full range of tools. However, it remains that the measures we included are relevant and widely utilized, affirming their significance for our analysis.

### Conclusion

In conclusion, no single questionnaire stands out as the “best” or most comprehensive. Each measure appeared to serve a different purpose rather than converging on unified constructs, making different tools suited for different use cases. Moving forward, it is recommended that questionnaires be developed with a clear framework, a transparent account of the construct’s origin and a focus on key aspects of the experience being measured. Future measures should adopt a more structured methodology to enhance measure development.

## Data Availability

Supplementary material data are available online at the Borealis data repository.

https://borealisdata.ca/dataverse/bilingualismmeasures/

## Funding, Disclosure and Data Availability Statement

This research was supported by the Natural Sciences and Engineering Research Council of Canada Discovery Grant (RGPIN-2019-06523) to M.M, Ontario Graduate Scholarship to KIL.

The authors have no conflicts of interest to disclose.

The data (supplementary materials) that support the findings of this study are openly available in Borealis at https://borealisdata.ca/dataverse/bilingualismmeasures/

## Footnotes

1 The Q-BEx offers a modular questionnaire with mandatory and optional questions depending on the user’s needs and interests. We specifically focused on the modules recommended to practitioners for its practicality and relevance for clinical use.

## Notes

### Competing Interest Statement

The authors have declared no competing interest.

### Funding Statement

This study was funded by the Natural Sciences and Engineering Research Council of Canada Discovery Grant (RGPIN-2019-06523) to M.M, Ontario Graduate Scholarship to KIL.

